# Evaluating Traditional, Deep Learning, and Subfield Methods for Automatically Segmenting the Hippocampus from MRI

**DOI:** 10.1101/2024.08.06.24311530

**Authors:** Sabrina Sghirripa, Gaurav Bhalerao, Ludovica Griffanti, Grace Gillis, Clare Mackay, Natalie Voets, Stephanie Wong, Mark Jenkinson, the Alzheimer’s Disease Neuroimaging Initiative

## Abstract

Given the relationship between hippocampal atrophy and cognitive impairment in various pathological conditions, hippocampus segmentation from MRI is an important task in neuroimaging. Manual segmentation, though considered the gold standard, is time-consuming and error-prone, leading to the development of numerous automatic segmentation methods. However, no study has yet independently compared the performance of traditional, deep learning-based, and hippocampal subfield segmentation methods within a single investigation. We evaluated nine automatic hippocampal segmentation methods (FreeSurfer, FastSurfer, FIRST, e2dhipseg, HippMapper, Hippodeep, FreeSurfer-Subfields, HippUnfold and HSF) across three datasets with manually segmented hippocampus labels. Performance metrics included overlap with manual labels, correlations between manual and automatic volumes, diagnostic group differentiation, and systematically located false positives and negatives. Most methods, especially deep learning-based ones, performed well on public datasets but showed more error and variability on unseen data. Many methods tended to over-segment, particularly at the anterior hippocampus border, but were able to distinguish between healthy controls, MCI, and dementia patients based on hippocampal volume. Our findings highlight the challenges in hippocampal segmentation from MRI and the need for more publicly accessible datasets with manual labels across diverse ages and pathological conditions.

**Key Messages:**

1. We evaluated nine automatic hippocampal segmentation methods, including traditional and deep learning-based approaches, across three datasets with manually segmented hippocampus labels.
2. While deep learning-based methods perform well on public datasets, they show more error and variability on unseen data that is more reflective of a clinical population.
3. More publicly accessible datasets with manual labels are required for automatic hippocampal segmentations to be accurate and reliable, particularly for clinical populations.

**Practitioner Points:**

1. Although deep learning based automatic hippocampal segmentation methods offer faster processing times—a requirement for translation to clinical practice—the lack of variance within training sets (such as sample demographics and scanner sequences) currently prevents transfer of learning to novel data, such as those acquired clinically.
2. More training data with varying demographics, scanner sequences and pathologies are required to adequately train deep learning methods to quickly, accurately and reliably segment the hippocampus for use in clinical practice.

## 1 Introduction

Using structural magnetic resonance imaging (MRI) to analyse the morphology of the hippocampus is crucial in both clinical and research neuroimaging. The correlation between hippocampal atrophy and cognitive impairment has long been established (Nadel & O’Keefe, 1978), implicating the hippocampus in various neurological and psychiatric conditions such as Alzheimer’s disease (AD) (Barnes et al., 2009), mild cognitive impairment (MCI) (Jack et al., 1999), epilepsy (Thom, 2014) and schizophrenia (Csernansky et al., 1998). For example, hippocampal atrophy stands as a well-established biomarker of AD, with volumetric measurements demonstrating the capability to distinguish between healthy older adults and patients at mild to advanced stages of the disease (Frisoni et al., 2008; Jack et al., 1997). In the interest of early detection and treatment of neurodegenerative disorders such as AD, it is unsurprising that extensive research effort has been directed towards developing efficient and accessible methods to accurately and reliably segment the hippocampus from MRI.

Manual segmentation of the hippocampus from MRI is widely regarded as the "gold standard" for volumetric measurement (Dill et al., 2015). However, manual segmentation is a time-consuming process, rendering it impractical for analysing large datasets. Likewise, issues with inter- and intra-rater reliability are prevalent, and results often vary significantly depending on the manual segmentation protocol employed (Boccardi et al., 2011). In response to these limitations, numerous automatic hippocampal segmentation methods have been proposed. Traditional methods of subcortical segmentation, such as FreeSurfer (Fischl, 2012) and FIRST (Patenaude et al., 2011), rely on model-based approaches. Each of these methods employ a Bayesian framework, where FreeSurfer uses deformable registration to determine subcortical labels, while FIRST uses active shape and appearance models. However, these methods, in practice, are time-consuming and resource intensive, making them less efficient for processing large datasets.

Recently, there has been a surge in publicly available, supervised deep-learning-based segmentation algorithms, offering shorter runtimes and heightened accuracy compared to traditional methods (Goubran et al., 2020; Henschel et al., 2020; Thyreau et al., 2018). For example, methods using convolutional neural networks (CNN) learn from existing hippocampus labels in a set of training data to identify and extract features to segment the hippocampus, with the intention of learning how to segment unseen images, measured using a separate test set. However, these methods face challenges due to the limited number of datasets containing manually labelled ground truth images for training. Moreover, the variance within training sets — such as sample demographics and scanner sequences — is quite limited, which underscores the importance of assessing the transfer of the learnt segmentation approach to novel datasets, and datasets that may more accurately reflect real-world, clinical populations.

Most studies introducing a new hippocampal segmentation technique benchmark the new technique against traditional methods such as FreeSurfer and FIRST, and other, newer techniques such as recently published deep learning-based methods (Goubran et al., 2020). Likewise, hippocampal segmentation methods that segment subfields (DeKraker et al., 2022; Poiret et al., 2023) benchmark across other subfield segmentation methods, so it is unclear how these methods compare to traditional sub-cortical segmentation and deep learning based methods that segment the whole hippocampus. Currently, no study has aimed to independently assess the performance of traditional, deep learning-based and hippocampal subfield segmentation methods, within a single investigation.

Our objective was to assess the performance of established and recent techniques for segmenting the hippocampus from T1-weighted MR images across three distinct datasets containing manual hippocampus labels. Broadly, our inclusion criteria encompassed segmentation methods that are publicly available, freely downloadable, and accept a single T1-weighted (T1w) image as input. For methods focusing on hippocampal subfields, we collapsed over subfields and solely evaluated the entire hippocampal mask. We were interested in a range of performance metrics including overlap with manual labels, correlations between manual and automatically segmented volumes, ability to separate diagnostic groups based on volume, and the location and number of systematically located false positives and false negatives within a method.

## 2 Method

### 2.1 Datasets

Each segmentation method was tested against manual labels in three datasets: ADNI HarP, MNI-HISUB25 and an in-house dataset of a subset of images collected by the Oxford Brain Health Clinic (OBHC). Table 1 presents the demographics from each dataset.

**Table 1.** Demographic informaHon and hippocampal volume from manual labels for ADNI HarP, MNIHISUB25 and Oxford Brain Health Clinic (OBHC) datasets.

|  | ADNI HarP |  |  | MNI-HISUB25 | OBHC |  |  |
| --- | --- | --- | --- | --- | --- | --- | --- |
|  | <u>AD</u> | <u>MCI</u> | <u>CN</u> | <u>CN</u> | <u>Dementia</u> | <u>MCI</u> | <u>NDRD</u> |
| **N** | 44 | 45 | 42 | 25 | 17 | 8 | 4 |
| **Age (years)** |  |  |  |  |  |  |  |
| *Mean (SD)* | 74 (8) | 75 (8) | 76 (7) | 31 (7) | 81 (7) | 79 (5) | 75 (6) |
| *Range* | 63 - 90 | 60 - 87 | 61 - 90 | 21 - 53 | 72 - 101 | 73 - 86 | 66 - 82 |
| **Sex** |  |  |  |  |  |  |  |
| *Female* | 23 (52%) | 19 (42%) | 22 (52%) | 13 (52%) | 7 (41%) | 5 (63%) | 3 (75%) |
| *Male* | 21 (48%) | 26 (58%) | 20 (48%) | 12 (48%) | 10 (59%) | 3 (38%) | 1 (25%) |
| **Hippocampal Volume (mm<sup>3</sup>)** |  |  |  |  |  |  |  |
| *Mean (SD)* | 2417 (530) | 2678 (471) | 3165 (518) | 4386 (246) | 2012 (669) | 2163 (479) | 2844 (554) |
| *Range* | 1054 - 3953 | 1794 - 3878 | 2007 - 5140 | 3823 - 5036 | 1050 - 3490 | 1249 - 2920 | 2193 - 3711 |

The HarP dataset (Boccardi, Bocchetta, Morency, et al., 2015) uses data from the Alzheimer’s disease Neuroimaging Initiative (ADNI) database (adni.loni.usc.edu), and consists of 135 T1-weighted MRI scans. The ADNI was launched in 2003 as a public-private partnership, led by Principal Investigator Michael W. Weiner, MD, and is primarily focused on investigating the progression of MCI and early AD. The dataset contains 1.5 T and 3 T-MRI scans acquired at a resolution of 1.0 x 1.0 x 1.2 mm with scanners from multiple MRI manufacturers (GE, Philips and Siemens). The images were acquired using an MP-RAGE sequence with parameters optimised for different scanners — for more details, see (Jack Jr. et al., 2008). Manual labels were created using the HarP protocol that is described in detail elsewhere (Boccardi, Bocchetta, Apostolova, et al., 2015), but briefly, a HarP segmentation includes the whole hippocampal head, body and tail, the alveus and whole subiculum based on the boundary with the entorhinal cortex. The dataset consists of 45 subjects with Alzheimer’s disease (AD), 46 mildly cognitively impaired subjects (MCI) and 44 older adult control subjects (CN).

The MNI-HISUB25 (Kulaga-Yoskovitz et al., 2015) dataset contains manual hippocampal subfield labels traced from T1 and T2 weighted images collected from 25 healthy subjects. Data were acquired on a 3T MRI using a 32 phased-array head coil. The manual protocol was guided by intensities and morphological characteristics of the of the hippocampal molecular layer and divided the hippocampus into 3 subregions: subicular complex, Cornu Ammonis (merged CA1, 2 and 3 regions) and CA-4-dentate gyrus.

The Oxford Brain Health Clinic (OBHC) is a joint clinical-research service for patients of the UK National Health Service (NHS) who have been referred to a memory clinic (O’Donoghue et al., 2023). At the OBHC, patients are offered high-quality assessments not routinely available, including a multimodal brain MRI scan acquired on a Siemens 3T Prisma scanner using a protocol matched with the UK Biobank imaging study (Griffanti et al., 2022; Miller et al., 2016). Patients are invited to participate in research and over 90% consented to their clinical data, including subsequent diagnosis, being used for research purposes, representing a real-world clinical dataset. The data is stored on the OBHC Research Database which was reviewed and approved by the South Central – Oxford C research ethics committee (SC/19/0404). Manual labels for the hippocampi were created for 29 subjects (17 patients who received a dementia diagnosis, 8 MCI patients, and 4 with no dementia-related diagnosis – NDRD).

### 2.2 Automatic Segmentation Methods

We evaluated the performance of 9 segmentation algorithms. To be included in our study, the algorithm was required to be freely and publicly available for download. There were 6 algorithms that performed hippocampus-only segmentations (Hippodeep, HippMapper, e2dhipseg, HippUnfold, HSF, FreeSurfer-Subfields), and 3 that created segmentations of a range of brain structures (FreeSurfer, FastSurfer, FIRST). For comprehensive information on each of the segmentation methods, please see the related publications. All algorithms were run using the default or recommended settings with a raw, full head T1w image as input, on either a MacBook Pro (Apple M2 Pro 2023, 16GB RAM), a machine with a NVIDIA GeForce GTX 1070, running Ubuntu V22.04 or on a dedicated cluster composed of CPU servers (16GB RAM per core) , GPU servers (K-series NVidia with CUDA) and Grid Engine queuing software.

<u>Hippodeep</u> (Thyreau et al., 2018) was run using the PyTorch version (https://github.com/bthyreau/hippodeep_pytorch) on a CPU (MacBook Pro or cluster). Hippodeep is based on a CNN trained on hippocampus outputs from FreeSurfer that were derived from 2500 images spanning 4 large cohort studies, as well as a small in-house sample of manually labelled ground truth images. The input to the algorithm was a T1w image, and the output of the algorithm was a probabilistic segmentation map, which was thresholded at 0.5 as recommended.

<u>HippMapp3r</u> (Goubran et al., 2020) version 0.1.1 was run using (https://github.com/AICONSlab/HippMapp3r/tree/master) on a GPU (GeForce GTX 1070) or via Singularity (cluster). HippMapper is based on a 3D CNN trained on manual segmentations from 259 older adults with a range of diagnoses leading to extensive atrophy, vascular disease and lesions. The input to the algorithm was a T1w image, which can either be whole or skull stripped. For these analyses, a whole T1w was provided. The output was a single file containing a binarised, bilateral hippocampal mask, which was then separated into a left and right hippocampus mask file.

<u>e2dhipseg</u> (Carmo et al., 2021) was run on a CPU (MacBook Pro or cluster) using the recommended affine registration option (FLIRT) (https://github.com/MICLab-Unicamp/e2dhipseg). The architecture of e2dhipseg consists of an ensemble of 2D CNNs which were trained on the ADNI HarP dataset and 190 images collected locally for an epilepsy study. The algorithm creates a set of segmentations that are then post-processed into a final hippocampal mask. The input to the algorithm was a T1w image, and the output was a single mask file containing both left and right hippocampal masks. The output mask was divided into a left and right hippocampus mask file, and then thresholded at 0.5 as recommended.

<u>HippUnfold</u> (DeKraker et al., 2022) version 1.3.0 is a BIDS app that was run using Docker or Singularity (cluster) (https://github.com/khanlab/hippunfold). HippUnfold uses CNNs and topological constraints to generate folded surfaces that correspond to an individual subject’s hippocampal morphology and was designed and trained with the Human Connectome Project 1200 young adult data release. The data is first gathered via snakebids before pre-processing, tissue class segmentation, post-processing and unfolding (see DeKraker et al., 2020). As this algorithm is a BIDS app, data from all datasets were first renamed and organised into the BIDS file structure format, before running HippUnfold using the default settings. The input is either a T1w image, a T2w image or both, and several output files are generated including hippocampal masks with subfields, surface metrics and warps. Here, HippUnfold was run with a T1w image as input, and the resulting hippocampal masks with subfields were binarised and combined into a whole hippocampus mask.

<u>Hippocampal Segmentation Factory (HSF)</u> version 4.0.0 was run on a GPU (MacBook Pro or cluster) (https://github.com/clementpoiret/HSF). The HSF pipeline includes brief pre-processing of raw T1w or T2w images and segmentation of hippocampal subfields through deep learning models trained on images from 12 datasets (411 subjects in total) with manual labels spanning across the chronological age range and multiple pathological groups. HSF was run using the default and/or recommended settings, including using the ROILoc package (https://github.com/clementpoiret/ROILoc) to centre and crop T1w images. The input was a T1w image and the output includes hippocampal masks with subfields, which were then binarised into a whole hippocampus mask.

<u>FIRST</u> (Patenaude et al., 2011) was run on a CPU (MacBook Pro or cluster). FIRST is a Bayesian appearance model-based tool that incorporates both shape and intensity information for segmentation, derived from a set of images from 336 subjects, each of which was manually segmented by the Center for Morphometric Analysis (CMA). FIRST was run using the *run_first_all* command specifying *L_Hipp* and *R_Hipp* as structures to segment. The output mask was then binarised using the recommended threshold.

<u>FreeSurfer</u> (Fischl, 2012) version 7.4.1 was used. FreeSurfer was also developed using 336 subjects each manually segmented by the CMA. The recon-all pipeline with default settings was run on all T1 images using parallel processing (computation time approximately 4 hours per subject on CPU). The steps in the recon-all pipeline are reported in detail elsewhere (Fischl et al., 2002). Hippocampal masks were extracted from the aseg.auto output and binarised.

<u>FreeSurfer Hippocampal Subfields and Nuclei of Amygdala</u> (Iglesias et al., 2015) (from now on referred to as “<u>FreeSurfer-Subfields</u>”) was developed using manual labels from *ex-* and *in-vivo* samples that were combined using a Bayesian inference based atlas building algorithm to form a single computational atlas. The data included 15 autopsy samples scanned at 0.13 mm isotropic resolution that were manually segmented into 13 different hippocampal structures, and a separate in-vivo dataset of T1w whole brain MRI (1 mm resolution) containing annotations for neighbouring structures such as the amygdala and cortex. FreeSurfer-Subfields requires a whole brain T1w image that has been processed with *recon-all* as input. The hippocampus component of the output masks (head, body, tail mask) was then binarised to create a whole hippocampus mask.

<u>FastSurfer</u> (Henschel et al., 2020) version 2.2.0 (https://github.com/Deep-MI/FastSurfer/) was run using Docker or Singularity (cluster). FastSurfer is a deep learning-based method that aims to present an alternative to FreeSurfer with a shorter run time. The FastSurferCNN architecture contains 3 CNNs operating on coronal, axial and sagittal 2D slices, and is trained on FreeSurfer parcellation following the Desikan–Killiany–Tourville protocol atlas. The seg_only function was used to generate subcortical segmentations from a T1w image, and from these, hippocampal masks were extracted and binarised.

For FreeSurfer, FreeSurfer-Subfields and FastSurfer, post-processing steps were required to allow the hippocampal masks to be compared with other segmentation algorithms. To do this, hippocampal masks were transferred back to the original image space using *mri_label2vol* and converted from mgz to NiFTi format using the function *mri_convert*. For these methods, hippocampal volumes were extracted from the relevant aseg.stats output file.

### 2.3 Validation metrics

Segmentation performance of each segmentation method was evaluated using common medical imaging segmentation metrics: Dice coefficient, 95^th^ percentile Hausdorff Distance and Pearson correlation between manual and automatically segmented volumes. Dice coefficients and Hausdorff distance were implemented using the seg-metrics Python package (Jia et al., 2024).

Dice coefficient measures the overlap between two binary sets. It ranges from 0 to 1, where 1 indicates a perfect overlap between the ground truth (manually segmented) and predicted (automatically segmented) masks. Hausdorff Distance was used as a metric to assess similarity in shape between the manual and automatic segmentations and is defined as the 95^th^ percentile of the distances between the closest points in an automatic segmentation mask and manual segmentation mask. HD95 is measured in millimetres, with a value of 0mm indicating perfect prediction.

### 2.4 Error maps

Average error maps for each segmentation method were also created to understand where each method was systematically incorrect in specific regions of the segmentation. False positive error maps were calculated by subtracting the manual mask from the automatic mask, using *fslmaths,* and keeping only positive values, while for false negatives the negative values were kept. Individual T1w images were linearly (*FLIRT*) and non-linearly (*FNIRT*) registered to MNI space, and the resulting warp fields were applied to transform individual false positive and false negative error maps into MNI space. The false negatives and false positives were then averaged and shown on the T1 1mm MNI template for visualisation.

### 2.5 Statistics

Statistical analyses and data visualisations were performed using R (R Version 4.3.1) and ggplot2 (Wickham, 2016).

Linear mixed effects models run using lme4 (Bates et al., 2015) were used to determine whether there were diagnostic group differences in hippocampal volume across segmentation methods. In these analyses, hippocampal volume was the outcome variable, segmentation method and diagnostic group were the fixed effects and subject was the random effect. Post-hoc pairwise *t*-tests were performed to explore group differences within segmentation methods, but due to the exploratory nature of the analyses, no multiple comparisons corrections were applied. In these tests, a *p*-value of less than 0.05 was considered statistically significant. Associations between manually and automatically segmented volumes were performed using Pearson’s correlation.

Data are presented as mean ± SD in text, mean (SD) in tables and mean ± SD in figures unless otherwise stated.

## 3 Results

### 3.1 ADNI HarP

#### 3.1.1 Segmentation Metrics

Hippmapper and e2dhipseg exhibited the highest Dice values across all conditions, with mean scores of 0.90 ± 0.03 and 0.89 ± 0.02 respectively. Hippodeep and FIRST demonstrated comparable performance, yielding mean Dice scores of 0.82 ± 0.03 and 0.80 ± 0.03 respectively (Figure 1A). The hippocampal subfield methods demonstrated similar performance based on mean Dice, but HippUnfold (0.75 ± 0.07) and HSF (0.74 ± 0.09) showed larger variance than FreeSurfer-Subfields (0.71 ± 0.04). FastSurfer (0.71 ± 0.04) and FreeSurfer (0.70 ± 0.05) demonstrated the poorest mean Dice scores, but relatively tight Dice distributions. Similarly, the 95^th^ percentile Hausdorff Distance was smallest for e2dhipseg (1.00 ± 0.04) and Hippmapper (1.01 ± 0.06), while HSF demonstrated the largest value (2.45 ± 2.47) (Figure 1B). Detailed results, including Dice coefficients and 95^th^ percentile Hausdorff Distance for each group and hemisphere, per segmentation method, are provided in Table 2.

**Figure 1.**
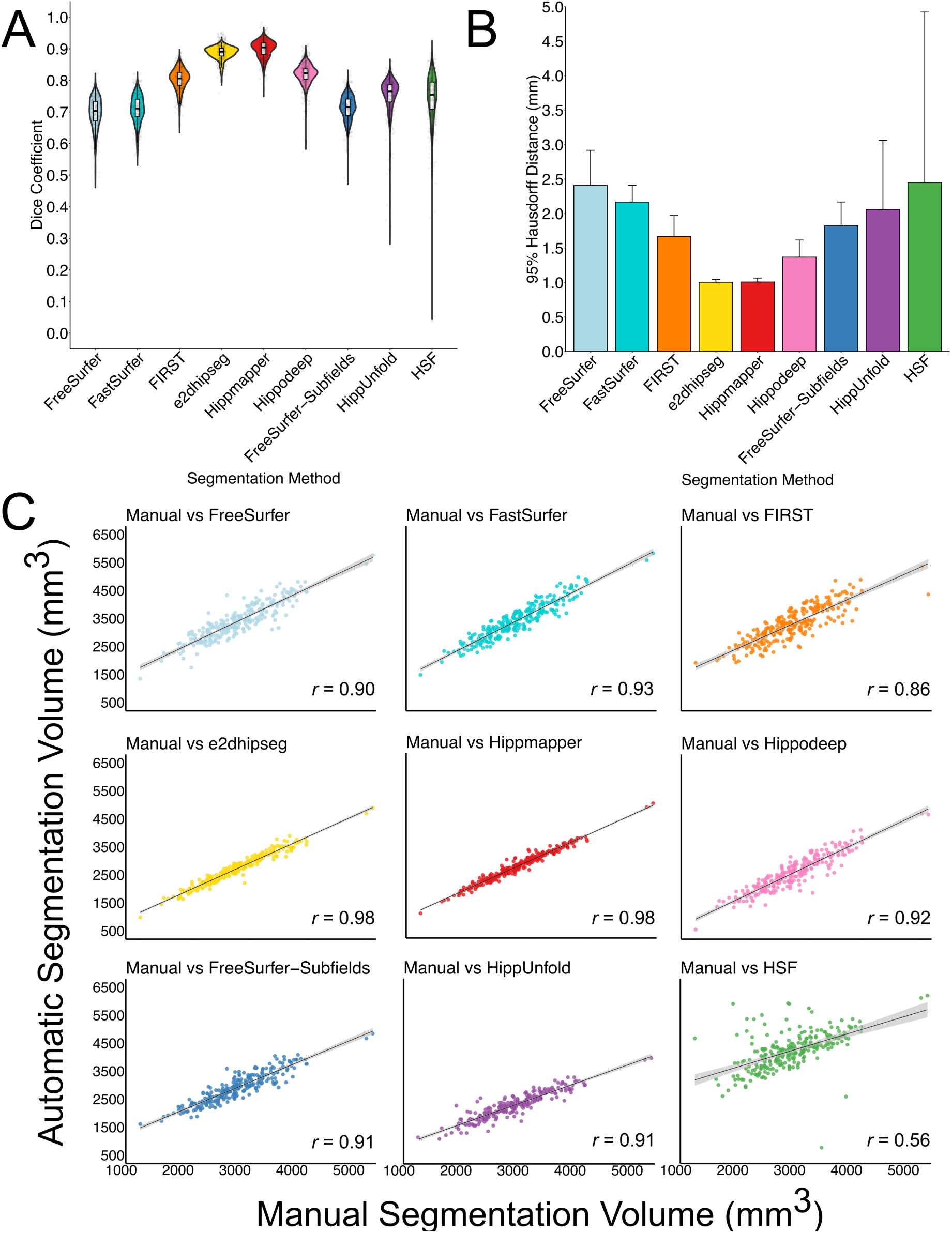
A) Dice coefficients and B) mean 95% Hausdorff Distance for automatic segmentation methods compared with manual hippocampal masks from the ADNI HarP dataset. C) Correlations between manually segmented and automatically segmented volumes for each segmentation method, averaged over diagnostic group and hemisphere.

**Table 2.** Mean volume, correlation coefficients, mean Dice coefficient and mean 95% Hausdorff distance across diagnostic groups and hemisphere for the ADNI HarP dataset.

EVALUATING HIPPOCAMPAL SEGMENTATION METHODS
|  | FreeSurfer | FastSurfer | FIRST | e2dhipseg | Hippmapper | Hippodeep | FreeSurfer-Subfields | HippUnfold | HSF |
| --- | --- | --- | --- | --- | --- | --- | --- | --- | --- |
| **Left Hemisphere Volume (mm<sup>3</sup>)** |  |  |  |  |  |  |  |  |  |
| AD | 3,019 (583) | 2,971 (562) | 2,879 (581) | 2,322 (488) | 2,373 (497) | 2,096 (547) | 2,537 (471) | 2,007 (434) | 4,021 (633) |
| MCI | 3,340 (508) | 3,362 (496) | 3,153 (467) | 2,616 (438) | 2,634 (430) | 2,454 (452) | 2,833 (451) | 2,232 (387) | 4,042 (557) |
| CN | 3,734 (590) | 3,841 (578) | 3,636 (521) | 3,076 (480) | 3,104 (481) | 2,922 (512) | 3,213 (489) | 2,582 (388) | 4,384 (805) |
| **Right Hemisphere Volume (mm<sup>3</sup>)** |  |  |  |  |  |  |  |  |  |
| AD | 3,083 (646) | 3,094 (620) | 3,041 (613) | 2,466 (537) | 2,520 (517) | 2,240 (614) | 2,649 (505) | 2,077 (448) | 4,236 (659) |
| MCI | 3,420 (542) | 3,407 (525) | 3,300 (535) | 2,701 (454) | 2,727 (446) | 2,549 (483) | 2,943 (487) | 2,283 (390) | 4,202 (544) |
| CN | 3,785 (550) | 3,887 (563) | 3,775 (534) | 3,129 (448) | 3,166 (465) | 3,044 (514) | 3,315 (432) | 2,599 (405) | 4,488 (589) |
| **Left Hemisphere Correlation with Manual Volume (r)** |  |  |  |  |  |  |  |  |  |
| AD | 0.81 | 0.92 | 0.88 | 0.97 | 0.98 | 0.89 | 0.90 | 0.84 | 0.43 |
| MCI | 0.85 | 0.91 | 0.82 | 0.98 | 0.97 | 0.89 | 0.85 | 0.93 | 0.82 |
| CN | 0.88 | 0.88 | 0.72 | 0.96 | 0.97 | 0.90 | 0.85 | 0.93 | 0.48 |
| **Right Hemisphere Correlation with Manual Volume (r)** |  |  |  |  |  |  |  |  |  |
| AD | 0.90 | 0.93 | 0.76 | 0.95 | 0.97 | 0.86 | 0.92 | 0.88 | 0.46 |
| MCI | 0.92 | 0.94 | 0.84 | 0.98 | 0.97 | 0.92 | 0.91 | 0.92 | 0.64 |
| CN | 0.89 | 0.89 | 0.82 | 0.95 | 0.98 | 0.90 | 0.88 | 0.90 | 0.48 |
| **Left Hemisphere Dice Coefficient** |  |  |  |  |  |  |  |  |  |
| AD | 0.67 (0.05) | 0.69 (0.04) | 0.79 (0.03) | 0.87 (0.02) | 0.89 (0.03) | 0.80 (0.03) | 0.69 (0.04) | 0.71 (0.08) | 0.69 (0.09) |
| MCI | 0.69 (0.04) | 0.70 (0.04) | 0.80 (0.03) | 0.89 (0.02) | 0.90 (0.02) | 0.82 (0.03) | 0.71 (0.04) | 0.76 (0.04) | 0.75 (0.05) |
| CN | 0.74 (0.03) | 0.74 (0.03) | 0.82 (0.03) | 0.90 (0.01) | 0.91 (0.02) | 0.84 (0.02) | 0.74 (0.03) | 0.78 (0.02) | 0.76 (0.12) |
| **Right Hemisphere Dice Coefficient** |  |  |  |  |  |  |  |  |  |
| AD | 0.67 (0.05) | 0.68 (0.04) | 0.79 (0.04) | 0.88 (0.02) | 0.89 (0.03) | 0.80 (0.04) | 0.69 (0.05) | 0.71 (0.10) | 0.70 (0.09) |
| MCI | 0.69 (0.04) | 0.70 (0.04) | 0.80 (0.03) | 0.89 (0.02) | 0.90 (0.03) | 0.82 (0.03) | 0.71 (0.04) | 0.75 (0.06) | 0.75 (0.06) |
| CN | 0.73 (0.02) | 0.74 (0.03) | 0.82 (0.03) | 0.90 (0.01) | 0.91 (0.02) | 0.83 (0.02) | 0.74 (0.02) | 0.78 (0.03) | 0.78 (0.06) |
| **Left Hemisphere 95% Hausdorff Distance (mm)** |  |  |  |  |  |  |  |  |  |
| AD | 2.77 (0.82) | 2.26 (0.32) | 1.62 (0.24) | 1.00 (0.00) | 1.00 (0.00) | 1.41 (0.21) | 1.95 (0.49) | 2.47 (1.41) | 2.69 (1.35) |
| MCI | 2.48 (0.46) | 2.25 (0.23) | 1.69 (0.27) | 1.00 (0.00) | 1.01 (0.06) | 1.38 (0.26) | 1.94 (0.32) | 1.89 (0.78) | 2.08 (0.43) |
| CN | 2.20 (0.20) | 2.11 (0.14) | 1.70 (0.37) | 1.00 (0.00) | 1.00 (0.00) | 1.30 (0.21) | 1.82 (0.29) | 1.67 (0.31) | 2.78 (5.02) |
| **Right Hemisphere 95% Hausdorff Distance (mm)** |  |  |  |  |  |  |  |  |  |
| AD | 2.46 (0.42) | 2.18 (0.19) | 1.69 (0.37) | 1.02 (0.09) | 1.02 (0.09) | 1.46 (0.34) | 1.79 (0.32) | 2.45 (1.25) | 2.66 (1.18) |
| MCI | 2.40 (0.41) | 2.18 (0.28) | 1.72 (0.29) | 1.00 (0.00) | 1.02 (0.09) | 1.37 (0.22) | 1.74 (0.28) | 2.06 (1.01) | 2.11 (0.69) |
| CN | 2.12 (0.20) | 2.02 (0.16) | 1.60 (0.25) | 1.01 (0.04) | 1.00 (0.00) | 1.30 (0.19) | 1.68 (0.23) | 1.78 (0.36) | 2.39 (3.03) |

Data are displayed as Mean (SD), except for when presenting Pearson correlation coefficients

#### 3.1.2 Volumes

Hippmapper and e2dhipseg exhibited the strongest correlation between manual and automatic volumes, consistently yielding correlation coefficients exceeding 0.95 across all conditions (Figure 1C). In contrast, HSF displayed the weakest correlation, with coefficients ranging from 0.43 to 0.82 across groups and hemispheres, indicating variable discrepancies between manual and automatic volume estimates, particularly for the AD group. Apart from HSF, the segmentation methods demonstrated relatively consistent performance across diagnostic groups and hemispheres (Table 2).

### 3.2 MNI-HISUB25

#### 3.2.1 Segmentation Metrics

On average, HSF (0.86 ± 0.016) and HippMapper (0.86 ± 0.020) achieved the highest Dice coefficients, followed by Hippodeep (0.85 ± 0.02) and FIRST (0.83 ± 0.021). HippUnfold (0.79 ± 0.03), FastSurfer (0.79 ± 0.02), FreeSurfer-Subfields (0.78 ± 0.03) and FreeSurfer (0.76 ± 0.03) performed comparably, while e2dhipseg (0.68 ± 0.12) yielded the poorest results (Figure 2A). Similarly, HippMapper exhibited the smallest 95^th^ percentile Hausdorff Distance (1.40 ± 0.17), followed by HSF (1.42 ± 0.10) and FreeSurfer-Subfields (1.52 ± 0.25). Consistent with the Dice value and distribution, e2dhipseg performed the poorest (4.94 ± 6.20) (Figure 2B). Overall, there was far less variability in performance between segmentation methods in this dataset compared to ADNI HarP and OBHC, likely due to the lack of patient groups. Segmentation metrics separated by hemisphere are presented in Table 3.

**Figure 2.**
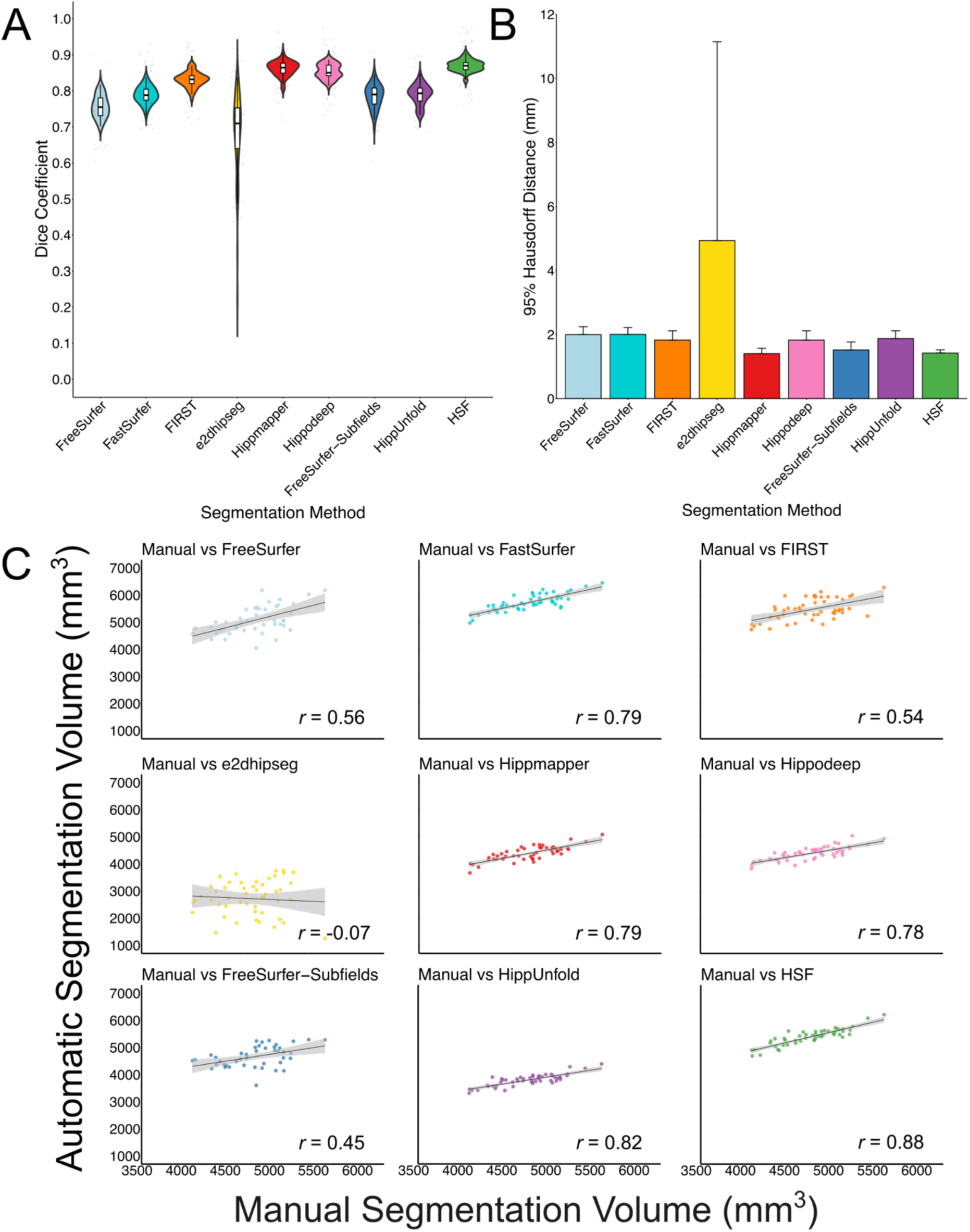
A) Dice coefficients and B) mean 95% Hausdorff Distance for automatic segmentation methods compared with manual hippocampal masks from the MNI-HISUB25 dataset. C) Correlations between manually segmented and automatically segmented volumes for each segmentation method, averaged over hemisphere.

**Table 3.** Mean volume, correlation coefficients, mean Dice coefficient and mean 95% Hausdorff distance across hemisphere for the MNI-HISUB25 dataset.

EVALUATING HIPPOCAMPAL SEGMENTATION METHODS
|  | FreeSurfer | FastSurfer | FIRST | e2dhipseg | Hippmapper | Hippodeep | FreeSurfer-Subfields | HippUnfold | HSF |
| --- | --- | --- | --- | --- | --- | --- | --- | --- | --- |
| **Left Hemisphere Volume (mm<sup>3</sup>)** |  |  |  |  |  |  |  |  |  |
| CN | 5,000 (475) | 5,701 (293) | 5,401 (368) | 2,590 (747) | 4,331 (263) | 4,354 (239) | 4,616 (391) | 3,767 (199) | 5,368 (286) |
| **Right Hemisphere Volume (mm<sup>3</sup>)** |  |  |  |  |  |  |  |  |  |
| CN | 5,144 (533) | 5,745 (333) | 5,522 (414) | 2,846 (625) | 4,443 (271) | 4,418 (254) | 4,708 (370) | 3,820 (235) | 5,416 (325) |
| **Left Hemisphere Correlation with Manual Volume (r)** |  |  |  |  |  |  |  |  |  |
| CN | 0.48 | 0.81 | 0.43 | -0.09 | 0.74 | 0.79 | 0.34 | 0.80 | 0.85 |
| **Right Hemisphere Correlation with Manual Volume (r)** |  |  |  |  |  |  |  |  |  |
| CN | 0.61 | 0.77 | 0.61 | -0.11 | 0.84 | 0.76 | 0.55 | 0.84 | 0.91 |
| **Left Hemisphere Dice Coefficient** |  |  |  |  |  |  |  |  |  |
| CN | 0.76 (0.03) | 0.79 (0.02) | 0.83 (0.02) | 0.66 (0.13) | 0.86 (0.02) | 0.85 (0.02) | 0.78 (0.03) | 0.80 (0.02) | 0.87 (0.01) |
| **Right Hemisphere Dice Coefficient** |  |  |  |  |  |  |  |  |  |
| CN | 0.76 (0.03) | 0.79 (0.03) | 0.83 (0.02) | 0.70 (0.11) | 0.86 (0.02) | 0.85 (0.02) | 0.79 (0.03) | 0.78 (0.03) | 0.87 (0.02) |
| **Left Hemisphere 95% Hausdorff Distance (mm)** |  |  |  |  |  |  |  |  |  |
| CN | 2.04 (0.17) | 2.05 (0.22) | 1.87 (0.33) | 6.22 (7.85) | 1.44 (0.14) | 1.87 (0.33) | 1.53 (0.18) | 1.85 (0.22) | 1.41 (0.11) |
| **Right Hemisphere 95% Hausdorff Distance (mm)** |  |  |  |  |  |  |  |  |  |
| CN | 1.95 (0.31) | 1.96 (0.21) | 1.78 (0.25) | 3.66 (3.70) | 1.37 (0.19) | 1.78 (0.25) | 1.51 (0.31) | 1.90 (0.27) | 1.44 (0.09) |

Data are displayed as Mean (SD), except for when presenting Pearson correlation coefficients

#### 3.2.2 Volumes

Correlations between manually and automatically segmented volumes notably weaken in the MNI-HISUB25 dataset compared to the ADNI HarP and OBHC datasets (Figure 2C). The strongest correlation between manual and automatic volumes was observed for HSF (*r* = 0.88), while no correlation was evident for e2dhipseg (*r* = -0.07). In this dataset, considerable differences in correlation coefficients between hemispheres were observed for certain methods, particularly in FIRST (right hemisphere: *r* = 0.61, left hemisphere: *r* = 0.43), FreeSurfer (right hemisphere: r = 0.61, left hemisphere: *r* = 0.48) and FreeSurfer-Subfields (right hemisphere: *r* = 0.55, left hemisphere: *r* = 0.34) (Table 3). Interestingly, there is a disconnect between Dice coefficients and volume correlations. Though FIRST was a strong performer based on Dice, the correlation between manual and automatic volumes was relatively weak (*r* = 0.54) compared to HSF (*r* = 0.88), HippMapper (*r* = 0.79) and Hippodeep (*r* = 0.78).

### 3.3 Oxford Brain Health Clinic

#### 3.3.1 Segmentation Metrics

On average, Hippodeep (0.76 ± 0.06) and FIRST (0.76 ± 0.12) exhibited the highest Dice coefficients, followed closely by FreeSurfer-Subfields (0.74 ± 0.06), although notable variation was observed in the Dice scores for FIRST. HippUnfold (0.72 ± 0.09), FastSurfer (0.71 ± 0.06), FreeSurfer (0.69 ± 0.07), and Hippmapper (0.69 ± 0.18) showed similar Dice values, with HippUnfold and Hippmapper displaying large distribution tails (Figure 3A). HSF (0.58 ± 0.15) and e2dhipseg (0.44 ± 0.26) were the poorest performers based on Dice, with both exhibiting substantial distribution tails attributed to failure rates. Conversely, the 95% Hausdorff Distance was smallest for HippUnfold (2.32 ± 1.09) and, as expected based on Dice, largest for e2dhipseg (6.90 ± 5.04) (Figure 3B). Additionally, in this dataset, FreeSurfer recon-all failed for two subjects who were removed from subsequent analysis. Overall, segmentation performance was considerably poorer in this dataset compared to ADNI HarP and MNI-HISUB25.

**Figure 3.**
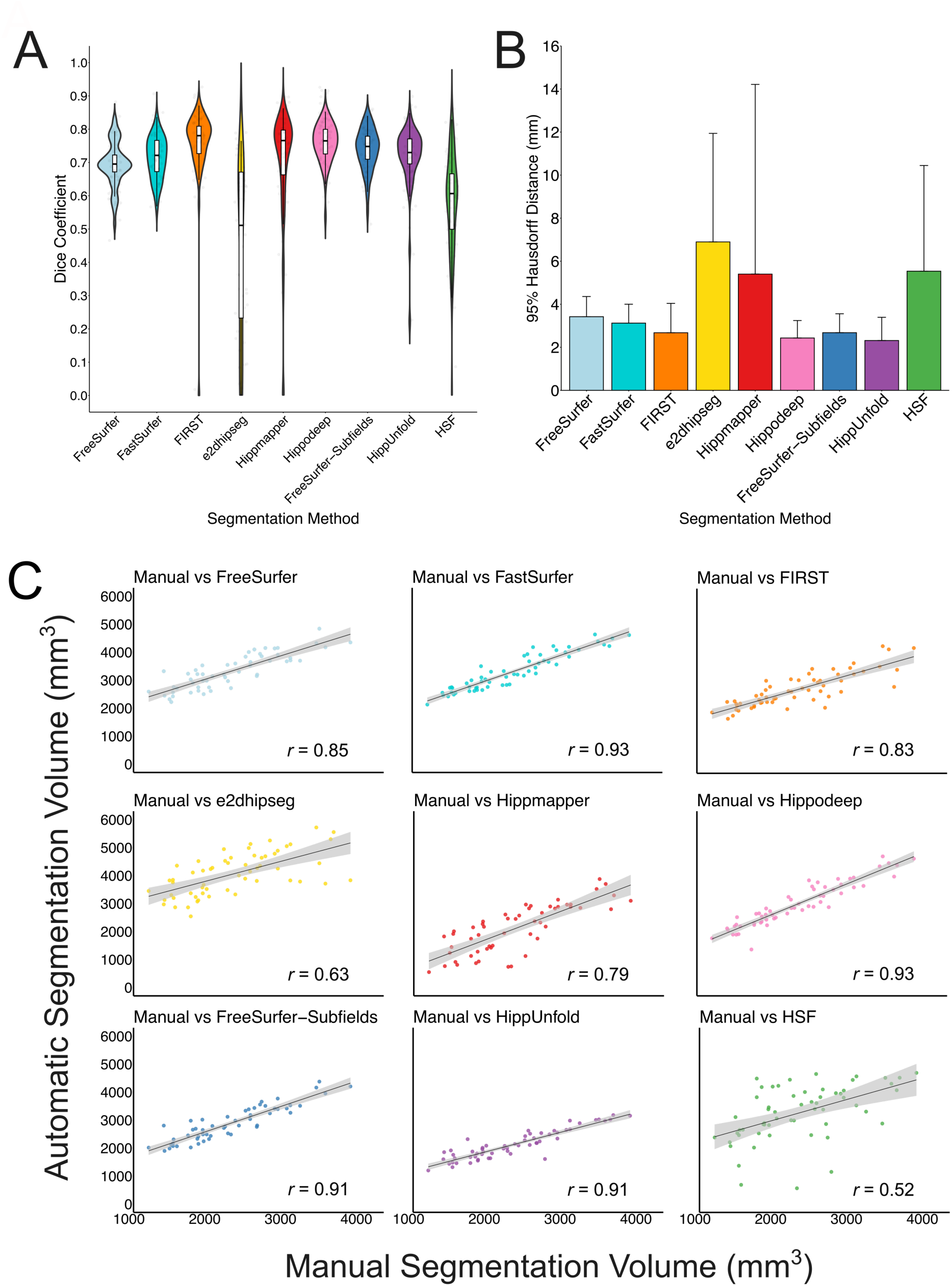
A) Dice coefficients and B) mean 95% Hausdorff Distance for automatic segmentation methods compared with manual hippocampal masks from the Oxford Brain Health Clinic dataset. C) Correlations between manually segmented and automatically segmented volumes for each segmentation method, averaged over diagnostic group and hemisphere.

#### 3.3.2 Volumes

Hippocampal volumes for each group and hemisphere, along with the correlation between manually segmented and automatically segmented volumes, are detailed in Table 4. FastSurfer and Hippodeep demonstrated the strongest correlation between manual and automatically segmented volumes on average (both *r* = 0.93) (Figure 3C), with both methods achieving correlation coefficients > 0.85 across hemisphere and diagnostic groups. HSF demonstrated the weakest correlation between manual and automatic volumes (r = 0.52), despite achieving correlation coefficients of 0.93 and 0.96 for the left and right hemisphere, respectively, in NDRD subjects. There is greater variation in correlation coefficients between diagnostic groups and hemispheres in the OBHC dataset compared to ADNI HarP. For example, FreeSurfer-Subfields demonstrated a correlation of *r* = 0.65 and *r* = 0.79 for subjects with an MCI diagnosis in the left and right hemisphere respectively, but all other correlations were > 0.89, while e2dhipseg performed particularly poorly in NDRD subjects compared with MCI and dementia groups (table 4).

**Table 4.**
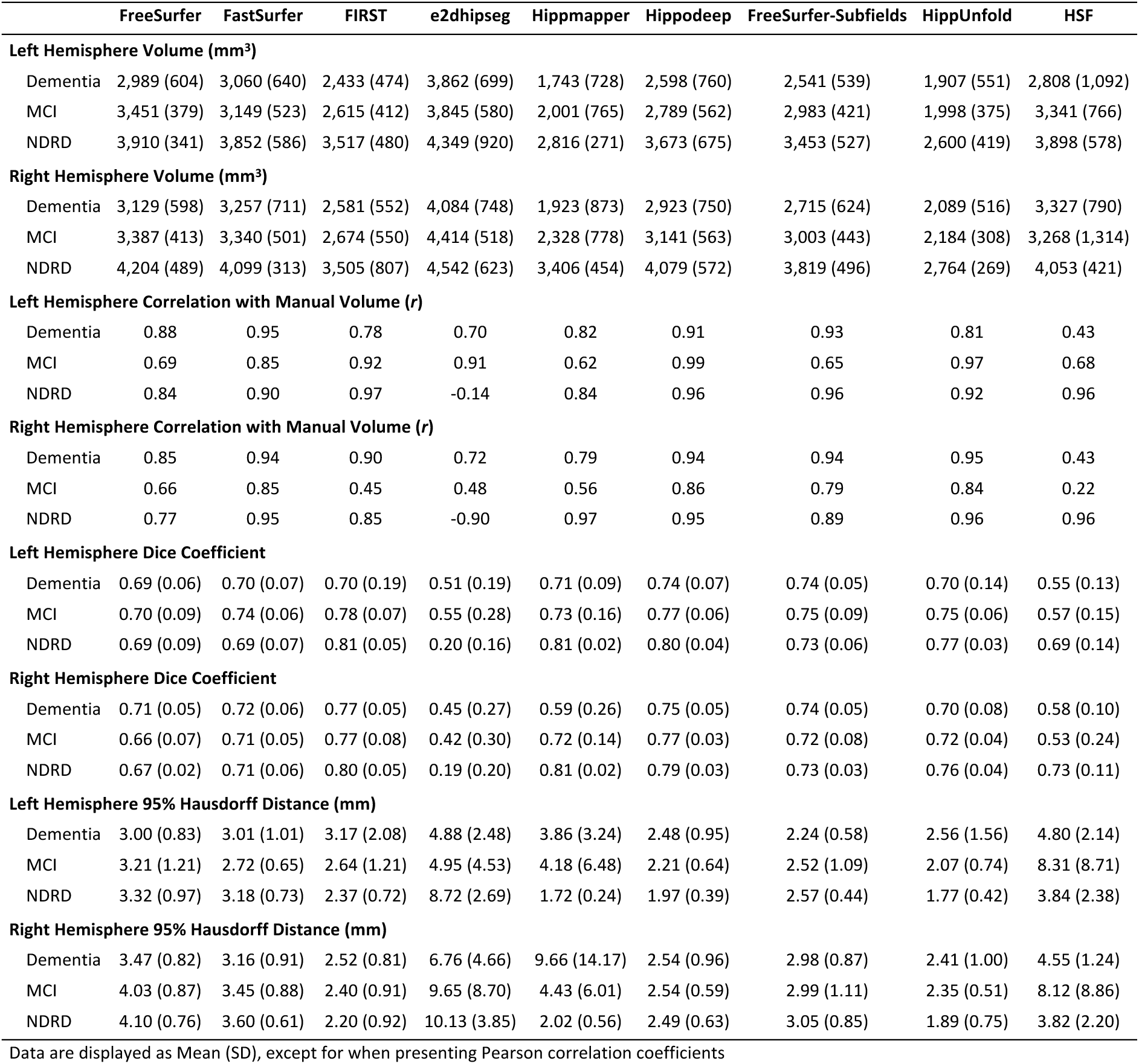
Mean volume, correlation coefficients, mean Dice coefficient and mean 95% Hausdorff distance across diagnostic groups and hemisphere for the Oxford Brain Health Clinic dataset.

### 3.4 Error Maps

To determine whether the segmentations were systematically failing in specific areas, heat maps detailing false positives and false negatives for each segmentation method in each dataset are shown in Figure 4.

**Figure 4.**
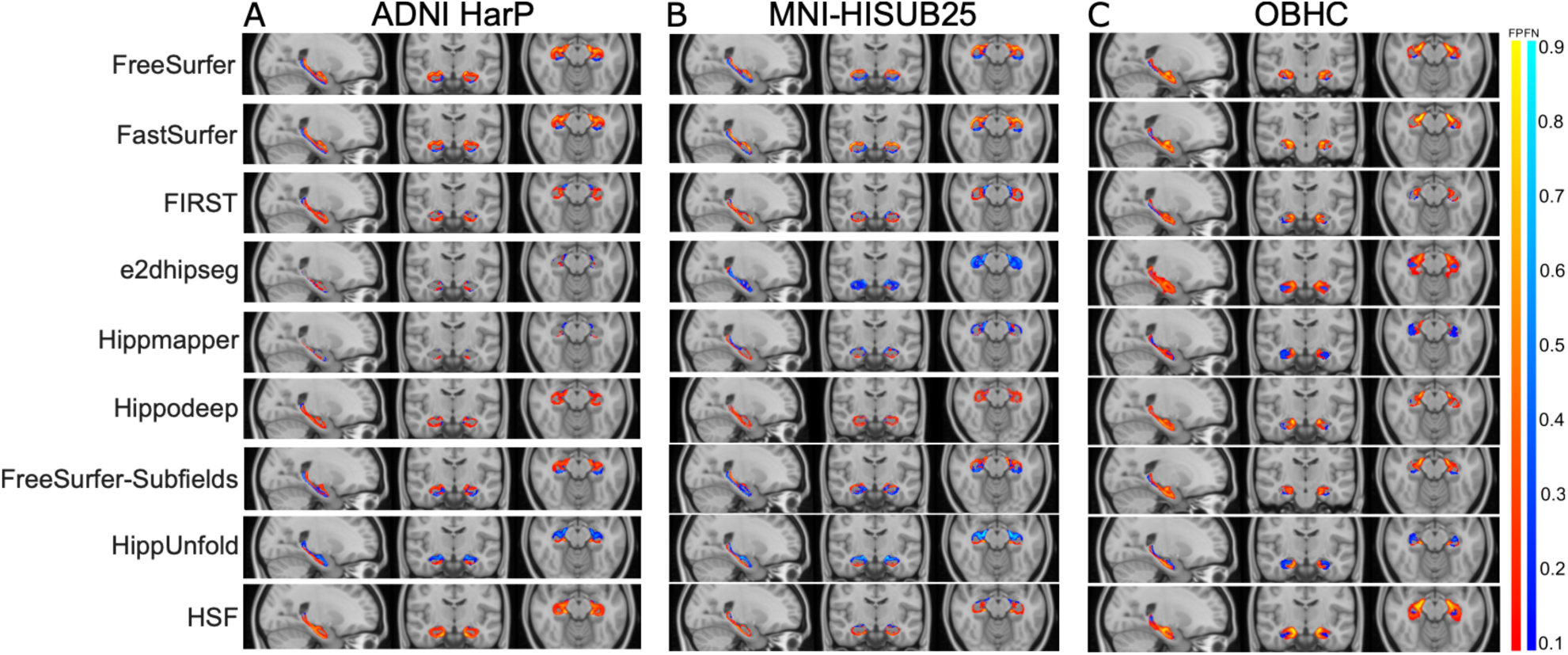
False positive (FP) (red-yellow) and false negative (FN) (blues) heat maps for each segmentation method projected onto the standard MNI152 T1 template image for A) ADNI HarP, B) MNI-HISUB25 and C) OBHC datasets. The colour bar represents the proportion of times a given voxel was incorrectly labelled in comparison to the manually segmented hippocampal mask.

In the ADNI HarP dataset, Hippmapper and e2dhipseg exhibited the lowest numbers of consistently located false negatives and false positives, while HippUnfold displayed relatively high numbers of consistently located false negatives. Overall, all segmentation methods, except for Hippmapper, e2dhipseg, and HippUnfold, primarily showed false positives in the error maps, suggesting a systematic over segmentation, particularly along the hippocampus-amygdala border. This tendency is most pronounced in the anterior hippocampal region in the axial slice (Figure 4A).

Figure 4B illustrates the systematic false positives and negatives for each segmentation method in the MNI-HISUB25 dataset. HSF and Hippmapper exhibit the fewest consistently located false positives and negatives, although all methods display false negatives along the superior-medial or inferior-medial border of the hippocampus. In line with its low Dice value and significant performance variation, e2dhipseg exhibits a high number of consistently located false negatives throughout the entire structure, indicating widespread under-segmentation.

Figure 4C depicts the systematic false positives and negatives for each segmentation method in the OBHC dataset. The incidence of consistently located false positives in the anterior region of the hippocampus is notably higher across many segmentation methods compared to the ADNI HarP and MNI-HISUB25 datasets. In agreement with the Dice values, Hippodeep, FIRST and FreeSurfer-Subfields demonstrate a relatively lower number of consistently located false negative voxels but exhibited systematic over-segmentation in the anterior portion of the hippocampus, while HippUnfold and Hippmapper tended to under-segment in the superior-medial hippocampal border. In line with having the lowest Dice values and significant performance variation, e2dhipseg shows false positives in all planes, including voxels outside of the hippocampus region incorrectly identified as hippocampal voxels.

### 3.5 Sensitivity to Diagnosis Groups

As assessing volumetric changes of the hippocampus is an important target in clinical contexts, we also sought to determine whether each segmentation method could detect changes in hippocampal volume between diagnostic groups in the ADNI HarP and OBHC datasets.

For the ADNI HarP dataset, a linear mixed effects model demonstrated a significant main effect of group (*F*2,128 = 24.55, *p* <0.001), segmentation method (*F*9,2431 = 1134.2, *p* <0.001) and a group by segmentation method interaction (*F*18,2431 = 7.43, *p* <0.001).

As a point of comparison, manual segmentation demonstrated that subjects with AD had smaller hippocampal volumes than control (*p*<0.001) and MCI subjects (*p*=0.04), and MCI subjects had smaller hippocampal volumes than controls (*p*<0.001). All automatic segmentation methods demonstrated smaller volumes in subjects diagnosed with AD compared with controls (all *p* <0.001). Likewise, all segmentation methods except for HSF (*p*=0.99) demonstrated lower volumes in subjects diagnosed as AD compared with MCI (all *p* < 0.04). Finally, all segmentation methods except HSF (*p*=0.05) demonstrated smaller hippocampal volumes in MCI subjects compared to controls (all *p*<0.005) (Figure 5A).

**Figure 5.**
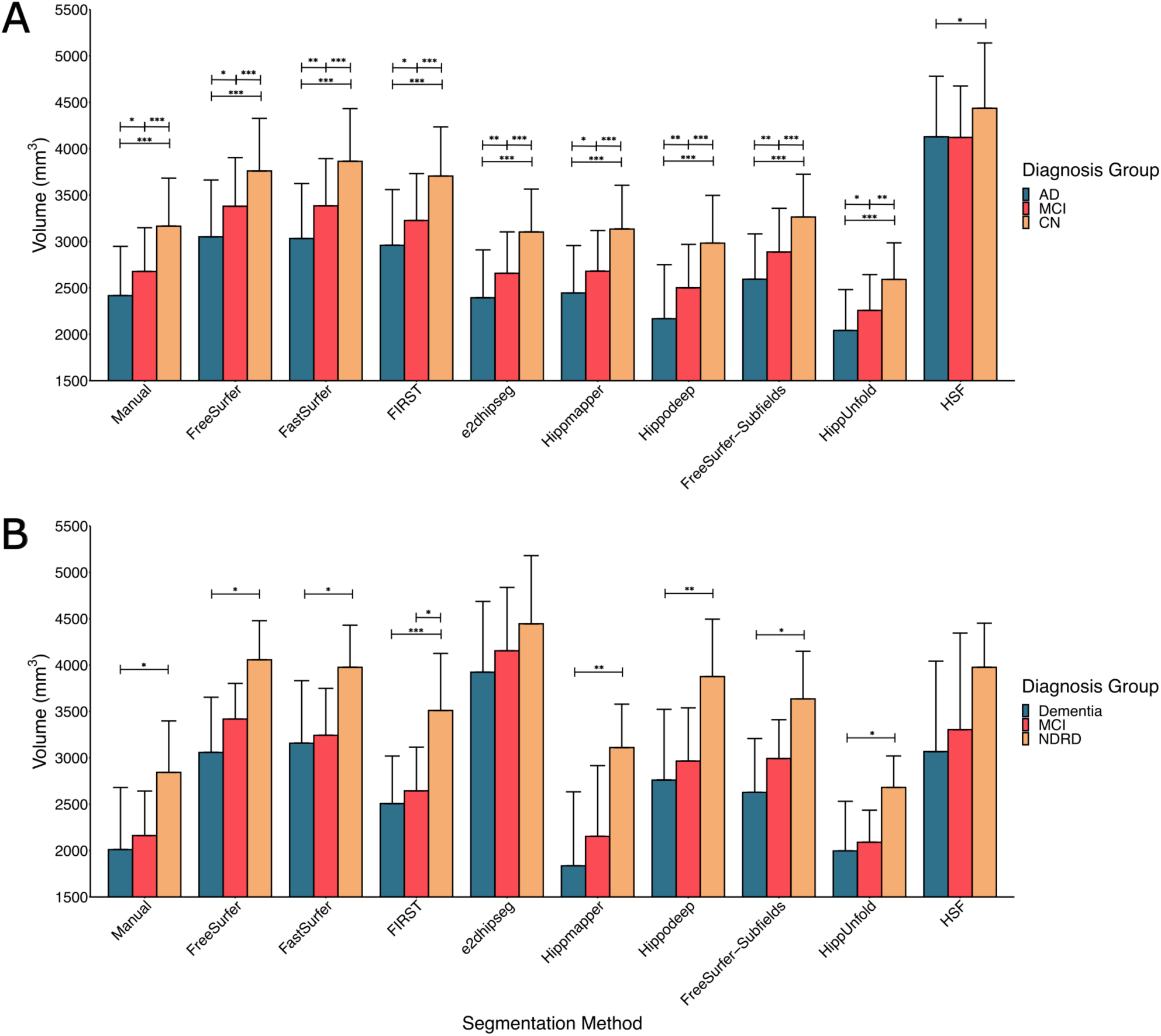
Mean volumes for each segmentation method across diagnostic groups in the A) ADNI HarP dataset and B) Oxford Brain Health Clinic dataset. *** *p* <0.001, ** *p* <0.01, * *p* <0.05

For the OBHC dataset, the model showed a significant main effect of group (*F*2,26 = 5.50, *p* = 0.01) and segmentation method (*F*9,511 = 74.4, *p* < 0.001), but no significant interaction between group and segmentation method (*F*18,511 = 0.86, *p* = 0.62). Though the group by segmentation method interaction was not statistically significant, comparisons between groups for each segmentation method were explored. For manual segmentation, subjects with a dementia diagnosis had smaller hippocampal volumes than those with no dementia-related diagnosis (*p* = 0.04). However, no significant differences were found in hippocampal volume between subjects with NDRD and MCI (*p* = 0.15), nor between subjects with MCI and dementia patients (*p* = 0.81). All segmentation methods (all *p* < 0.04) except e2dhipseg and HSF found smaller hippocampal volumes for subjects with a dementia diagnosis compared with NDRD (e2dhipseg: *p* = 0.36, HSF: *p* = 0.11). Only FIRST demonstrated a difference in volume between NDRD and subjects with MCI (*p* = 0.01), while no segmentation methods detected a difference in volume between subjects with a dementia or MCI diagnosis (all *p* > 0.24) (figure 5B).

## 4 Discussion

Here, we investigated the performance of nine publicly available tools that segment the hippocampus on three datasets with manual labels: ADNI HarP, MNI-HISUB25 and a real-world memory clinic dataset from the OBHC. Briefly, we found that most segmentation methods: 1) performed consistently and accurately on the two publicly available datasets but were more prone to error and variability on an unseen, clinical dataset, 2) are likely to systematically over-segment the hippocampus, particularly at the anterior hippocampus border, and 3) can delineate between healthy controls, subjects with a diagnosis of MCI and subjects with a dementia diagnosis based on hippocampal volume.

In evaluating the performance of each of the segmentation methods, we were interested in both accuracy relative to manual labels and reliability. Hippmapper and e2dhipseg excelled on the ADNI HarP dataset, showing near perfect Dice values, Hausdorff Distances and volume correlations.

However, HSF was the poorest performer, showing a large Dice distribution, weak correlations with manual volumes, and an inability to distinguish diagnostic groups based on volume. In the MNI-HISUB25 dataset however, HSF was the best performer, exhibiting high and consistent Dice values and strong correlations with manual volumes. Traditional methods maintained consistent performance across the ADNI HarP and MNI-HISUB25 datasets, although correlations with manual volumes were weaker compared to the deep learning-based methods in MNI-HISUB25.

However, performance on the OBHC dataset was generally worse across all methods. This is likely because the OBHC dataset has different demographic and clinical features than the other datasets. Being an unselected sample of memory clinic patients, the average age is older than ADNI and most dementia research datasets, and there is higher amount of atrophy and vascular pathology (Griffanti et al., 2022). Moreover, due to minimal exclusion criteria (related to MR-safety or being too frail to travel to the assessment centre – see O’Donoghue et al., 2023 for details), it is a more representative sample of real-world patients, but also likely more heterogeneous. If the goal is to make automated measures available in clinical practice, it is important to evaluate the performance of tools in such clinical samples. In this dataset, FIRST, e2dhipseg, Hippmapper, HippUnfold, and HSF exhibited lower mean Dice coefficients and worse tail distributions than in the other datasets, suggesting higher segmentation failure rates. However, Hippodeep, FastSurfer and FreeSurfer-Subfields performed relatively well on the OBHC data, with strong correlations with manual volumes and better accuracy and consistency than other methods as measured by Dice distributions, potentially suggesting better performance in data collected in real-world clinical settings and populations beyond typical research samples.

It is important to note instances where focusing solely on specific datasets or metrics would lead to arriving at different conclusions. For example, assessing only the ADNI HarP data would suggest e2dhipseg and Hippmapper as top performers. Results from the MNI-HISUB25 dataset would provide evidence against the efficacy of e2dhipseg, but further evidence for Hippmapper. However, the pronounced distribution tail of Dice values in the OBHC dataset would raise concerns about the reliability and failure rates of Hippmapper, despite its strong performance elsewhere. Similarly, in the MNI-HISUB25 dataset, although HSF demonstrated high Dice values, low variability, and strong correlations with manual volumes, its relatively poor performance on ADNI and OBHC datasets raises concerns about transferability. The development and validation methodology used by some of the deep learning segmentation methods can provide some insight into their performance. For example, Hippmapper (Goubran et al., 2020) and e2dhipseg (Carmo et al., 2021) were developed and validated on the ADNI HarP dataset, while HSF was trained on MNI-HISUB25 (Poiret et al., 2023), explaining the excellent performance on their respective seen datasets and poorer performance elsewhere. The diminished performance of most deep learning methods on unseen data emerges as a significant finding of this study.

Not all deep-learning-based methods, however, exhibited inconsistency across datasets. Hippodeep demonstrated strong performance across all segmentation metrics in all evaluated datasets, with high Dice values, tightly distributed Dice and Hausdorff Distance measures, and robust correlations with manual volumes. Likewise, although slightly underperforming compared with other methods based on mean Dice values, FastSurfer consistently demonstrated relatively tight Dice distributions and strong correlations with manual volumes across all datasets. The consistent performance of these methods can be attributed to training methods that allowed for larger and diverse datasets to be used in the development of these tools. FastSurfer implements a CNN that is trained on labels derived from FreeSurfer cortical and subcortical segmentation (Henschel et al., 2020), while Hippodeep was trained in part, on hippocampal labels derived from FreeSurfer (Thyreau et al., 2018). By reducing the reliance on manually labelled data, both methods accessed larger training datasets that spanned different ages, disease groups, scanner types, scanner vendors and field strengths, which likely explains the better performance on unseen data here.

In comparison, the traditional methods of FreeSurfer and FIRST, while slightly underperforming compared to deep learning-based methods in the ADNI and MNI-HISUB25 datasets, demonstrate relatively consistent performance across all datasets when considering mean Dice values, mean 95^th^ percentile Hausdorff distance, Dice distribution, and correlation with manual volumes. Although FIRST and FreeSurfer showed relatively weaker correlations with manual volumes in the MNI-HISUB25 dataset, and FIRST exhibited a large Dice distribution tail in the OBHC data, their performance across methods was generally moderate to strong, although FIRST outperformed FreeSurfer in both ADNI HarP and MNI-HISUB25. The consistency of traditional segmentation methods compared with deep learning-based methods highlights that a lack of manual label sources is a notable limitation of the hippocampal segmentation field. The shortage of manual labels that span the entire chronological age range and pathological conditions results in new methods being repeatedly trained on similar data, limiting generalisability even when the unseen data matches the demographic characteristics of common training datasets, such as ADNI HarP.

Another finding of this study is the consistent over-segmentation in the anterior hippocampal region among most segmentation methods, unless they perform exceptionally well (e.g., e2dhipseg and Hippmapper on ADNI HarP) or poorly (e.g., e2dhipseg on MNI-HISUB25). Even the most reliably performing methods, such as Hippodeep, exhibit systematic over-segmentation at the anterior hippocampus border, indicating challenges in delineating the boundaries between the hippocampus and amygdala. This difficulty is expected given the lack of visible landmarks to reliably demarcate regions within the medial temporal lobe, particularly between the borders of the hippocampus and amygdala, CA subregions, subiculum, and entorhinal cortex (Amunts et al., 2005). Additionally, consistently located false negatives, although less frequent, were seen at the medial and posterior borders of the hippocampus. This region is challenging to segment, as many manual segmentation protocols rely on the appearance of non-hippocampal structures to assist in defining hippocampal borders (Konrad et al., 2009). As cytoarchitecture is not visible in MR images, it is unsurprising that accurately and reliably segmenting the hippocampus both manually and automatically remains an ongoing challenge in the neuroimaging field.

A caveat to consider is that all three datasets in this study were labelled using different manual labelling methods, potentially influencing the comparative results across datasets and segmentation methods (Frisoni & Jack, 2011). For example, while the ADNI-HarP data was labelled using the extensively tested HarP segmentation protocol (Frisoni & Jack, 2015), the MNI-HISUB25 dataset was labelled with the intention to capture 3 broad regions (subiculum, CA1-3 and CA4-DG) (Kulaga-Yoskovitz et al., 2015). The automatic segmentation methods that were not trained on subfield data showed larger segmented volumes and weak correlations between manual and automatic volumes in the MNI-HISUB25 dataset despite the sample only containing healthy controls, which may indicate differences resulting from manual labelling methods. However, the consistent pattern of false negatives and false positives observed across segmentation methods and datasets may indicate that regardless of manual labelling protocol, automatic segmentation methods systematically fail in similar ways.

Selecting the most appropriate hippocampal segmentation method for a given analysis should be guided by research aims and available resources for processing. Based on the data presented, Hippodeep emerges as particularly attractive for solely segmenting the hippocampus, offering high similarity to manual masks based on Dice, strong correlations with manual volumes, the ability to detect group differences based on volume, and efficient processing times (i.e. processing a single participant in under 30 seconds on a CPU). If whole brain segmentation is of interest, FastSurfer presents a viable, computationally inexpensive alternative to FreeSurfer (i.e. performing whole brain segmentation in approximately 5 minutes on a GPU or 7 minutes on a CPU, compared with 4+ hours for FreeSurfer), demonstrating improved performance over FreeSurfer in all datasets. Although FIRST demonstrates higher mean Dice values than FastSurfer in all datasets, FastSurfer demonstrates stronger correlations with manual volumes, but both methods are sensitive to diagnostic group differences. However, if only ADNI data is of interest then there are other methods that could perform better. Alternatively, if hippocampal subfields are of interest, from the methods tested here, the combination of FreeSurfer and FreeSurfer-Subfields is likely the most reliable option, followed closely by HippUnfold, which also provides additional output such as surface data and Laplacian fields. However, both methods are computationally intensive and may require high-performance computing resources for large datasets, which may not be an option in all use cases. For studies investigating group differences in hippocampal volume, most of the segmentation methods evaluated in this study are suitable, even for subtle volume changes such as those between MCI and AD or MCI and control groups. Therefore, the choice of method should also consider other factors such as computational resources and compatibility with study objectives.

One limitation of this study is the relatively small sample size of the OBHC dataset, with a very small number of participants without a dementia-related diagnosis. However, this limitation is not unique to our study but rather reflects the broader challenge in the field of manual label availability. Moreover, the NDRD group in the OBHC dataset cannot be considered a healthy control group in a similar way as other datasets, as all OBHC participants were referred for a memory clinic appointment. We used as our third diagnostic group those patients who did not receive a diagnosis of MCI or dementia, but they may have received other mental-health diagnoses or no diagnosis. However, this group is more typical of a general clinical population. Another limitation is that we did not aim to optimise the automatic segmentation methods evaluated here; instead, we used default or recommended settings in all cases. While methods with adjustable parameters or additional input information (e.g., T2w or brain-extracted images) may show improved performance when these are provided, it was beyond the scope of this study to explore the optimisation of each method. Finally, it was also beyond the scope of this study to evaluate the performance of automatic segmentation methods longitudinally, but this is a natural extension of this work that we will focus on in the future.

In this study, we assessed the performance of nine automatic hippocampal segmentation methods across three datasets with manual labels. While it is challenging to provide a single, definitive recommendation for the most valid method(s) based on this investigation, our findings underscore the ongoing challenge of hippocampal segmentation from MR images within the neuroimaging field. As the field moves towards deep-learning-based segmentation, future efforts should prioritise increasing the availability of publicly accessible manual labels covering a wide range of ages and pathological conditions. This would facilitate adequate training of segmentation methods and enhance their generalisability, both cross-sectionally and longitudinally.

## Acknowledgments

We are grateful to the operations team of the OBHC (see https://www.psych.ox.ac.uk/research/translational-neuroimaging-group/team/oxford-brain-health-clinic)

## Data Availability Statement

The OBHC dataset will be available via the Dementias Platform UK (https://portal.dementiasplatform.uk/CohortDirectory) and access will be granted through an application process, reviewed by the OBHC Data Access Group. Data will continue to be released in batches as the OBHC progresses to minimise the risk of participant identification.

## Ethics Approval Statement

The storage of data on the OBHC Research Database was reviewed and approved by the South Central – Oxford C research ethics committee (SC/19/0404)

